# Molnupiravir’s real-world effectiveness in COVID-19 non-hospitalized patients at high risk of severe disease: a single-center study

**DOI:** 10.1101/2023.03.03.23286761

**Authors:** I Gmizic, N Todorovic, M Sabanovic, N Kekic, N Boskovic, I Milosevic, G Stevanovic

## Abstract

**Objective:** To assess the real-world effectiveness of molnupiravir (MOL) in reducing the need for hospitalization in at-risk, non-hospitalized patients with confirmed COVID-19.

**Methods:** A single-center, non-randomized, observational retrospective study of non-hospitalized patients with confirmed COVID-19 treated at the Clinic for Infectious and Tropical Diseases, University Clinical Center in Belgrade, Serbia.

**Results:** Between December 15^th^, 2021, and February 15th, 2022, 320 patients were eligible for inclusion in the study. Of these, 165 received treatment with molnupiravir (51.6%), while both groups were similar in gender and age distribution. The treatment group had a higher proportion of vaccination (75.2% vs. 51%, p<0.001) but was similar to the control group in terms of multiple comorbidity predomination (65.5% vs. 65.2%, p 0.956). The majority of patients who received MOL didn’t require hospitalization (92.7 vs. 24.5%, p<0.001) and needed oxygen supplementation less frequently than those in the control group (0.6% vs. 31%, p<0.001). During the follow-up period of 12.12±3.5 days, none of the patients on MOL were admitted to the Intensive Care Unit (vs. 10.3%, p<0.001). Molnupiravir significantly reduced the risk of hospitalization by 97.9% (HR 0.021, p<0.001).

**Conclussion:** Our study has proven the effectiveness of molnupiravir in preventing hospitalization in a population at risk for developing severe forms of COVID-19.

## INTRODUCTION

The World Health Organization (WHO) declared coronavirus disease 2019 (COVID-19), caused by SARS-CoV-2, a pandemic at the beginning of March 2020 [1]. The clinical appearance varied by severity, from mild to severe, and led to a substantial increase in morbidity and mortality worldwide. Many patients, especially the elderly and those with preexisting illnesses (e.g., obesity, diabetes mellitus, and major heart problems), required hospitalization, and many have died as a result of respiratory failure, shock, and multiple organ failure [2, 3, 4]. Virus genetic diversity presented an additional challenge, as successive variants differed not only in infectivity and pathogenicity but also in antiviral medication effectiveness. There has been an urgent need for effective forms of therapy to reduce the global health burden since the outbreak. Many clinical trials have shown that treatment should begin as soon as possible after the onset of symptoms and that it should ideally be readily available and easily administered by the patients themselves [5, 6].

Because it takes time to develop a new medicine, researchers have focused their efforts on repurposing existing pharmaceuticals for new applications. Molnupiravir (MOL), an antiviral medication that was initially studied in preclinical studies with influenza, has risen to prominence [7, 8]. MOL is a polymerase inhibitor prodrug that acts as a synthetic nucleoside; it is orally administered, and it’s used to treat COVID-19 and facilitates therapy in the out-of-hospital setting [9, 10]. MOL was evaluated in several phase 3 trials with the specific goal of assessing its effectiveness in at-risk, non-hospitalized adults who had COVID-19 symptoms for less than 5 days [11]. Based on the phase 2/3 MOVe-OUT clinical study in non-hospitalized patients with SARS-CoV-2 infection, the Food and Drug Administration (FDA) approved MOL for emergency use in the treatment of mild to moderate SARS-CoV-2 illness in December 2021 [12]. This study showed that early treatment with MOL reduces the risk of hospitalization or death in at-risk, unvaccinated adults with COVID-19 [12]. Our study aimed to assess the real-world effectiveness of MOL in reducing the need for hospitalization in at-risk, non-hospitalized patients with confirmed COVID-19.

## PATIENTS AND METHODS

In this single-center, non-randomized, observational retrospective study, we included data extracted from medical charts in the electronic medical database Heliant (Health information system) for non-hospitalized patients treated at the Clinic for Infectious and Tropical Diseases, University Clinical Center in Belgrade, Serbia. We included patients treated between December 15, 2021, and February 15, 2022, which is considered the period of dominance of the Omicron SARS-CoV-2 variant in Serbia. The decision about the treatment regimen was taken entirely by the treating physician, concerning current knowledge and recommendations of the National Protocol of Serbia for COVID-19. All the patients were diagnosed with COVID-19 based on positive results of the real-time reverse transcriptase polymerase chain reaction (RT-PCR) or an antigen test from the nasopharyngeal swab specimen. All the patients were non-hospitalized adults aged 18 years with mild or moderate COVID-19 and with associated risk factors for the development of severe illness from COVID-19 (age >60 years; active cancer; chronic kidney disease; chronic obstructive pulmonary disease; obesity, defined by a body mass index [the weight in kilograms divided by the square of the height in meters] of 30; serious heart conditions [heart failure, coronary artery disease, or cardiomyopathies]; or diabetes mellitus). Mild or moderate illness was determined on the basis of definitions adapted from World Health Organization (WHO) guidance [1]. In a “treatment group,” we included patients who started MOL in the first 5 days after the onset of symptoms. Molnupiravir was administered orally twice daily at 800 mg for 5 days according to the National Protocol of Serbia for COVID-19.

The “control group” included patients who didn’t take MOL due to multiple reasons: the medicine wasn’t available, they refused to take it, or they were diagnosed after the 5^th^ day of their symptoms. Patients for the control group were collected simultaneously and in such a way that they corresponded to the clinical criteria for the use of molnupiravir and that they had a similar distribution in terms of gender and age. Age, gender, symptoms with duration, risk factors (advance age, active malignancy, chronic kidney disease (CKD), chronic obstructive pulmonary disease (COPD), obesity, a heart condition, diabetes mellitus, autoimmune disease), use of other non-antiviral medications against COVID-19, vaccination status reported by patients, and time since the last vaccination were all included in the baseline data. We included laboratory measurements taken at the first and last examinations, including: C-reactive protein (CRP), white blood cells (WBC), platelets, d-dimer, aspartate transaminase (AST), and alanine transaminase (ALT). The primary effectiveness end point was the incidence of hospitalization for any cause (defined as 24 hours of acute care in a hospital or any similar facility). Patients were followed-up for 25 days or until the day of hospitalization. Moreover, the final outcome was analyzed using ordinal scale categories as follows: 0) unhospitalized; 1) hospitalized, requiring no oxygen supplementation but requiring medical care; 2) hospitalized, requiring normal oxygen supplementation; 3) hospitalized, on non-invasive ventilation with high-flow oxygen equipment; 4) hospitalized, on invasive mechanical ventilation.

## STATISTICAL ANALYSIS

Continuous variables with normal distribution of the data are presented as mean value ± standard deviation. The normal distribution of the data was checked using Kolmogorov-Smirnov and Shapiro-Wilk tests. Categorical variables are presented as the frequencies (percentage). For continuous variables, intergroup differences were tested using 2-sided t test while the χ_2_ or Fisher exact test was used to compare the distribution of categorical variables among groups. Univariate Cox regression model was used to identify the candidate predictors of the hospitalization due to COVID-19 while the independent predictors were analysed using the multivariate Cox regression analyses. P values were 2-sided and a *p* value of < .05 defined statistical significance. The statistical analyses was performed using the IBM SPSS software v21.

## RESULTS

Our study population included 165 patients while 155 patients were in the control group. Demografic and clinical characteristics of all patients are shown in Table 1. The study groups were similar in terms of sex and age, with as many as 62.4% of patients treated with MOL and 69.7% of the untreated group over 60 years of age. Observing the risk factors, the study group had statistically significantly more patients with heart diseases and active malignancy, while the study group had more chronic kidney patients and asthmatics. There was a significant difference in vaccination status between groups (75.2% vs. 51%, p<0.001), with the treatment group receiving three doses of vaccine (vs. one dose in the control group, p<0.001) 2.1–2.8 months prior to the onset of symptoms (vs. 0.97–1.9 months in the control group, p<0.001). The main difference between groups in previous COVID-19 treatment was that 56.1% of patients in the control group took antibiotics (vs. 9.1%, p<0.001), while 83% of patients in the treatment group took symptomatic treatment (vs. 40.6%, p<0.001). The prevalence of symptoms was pretty similar between groups, but cough and dyspnea were more present in the control group (65.5 vs. 80%, p <0.004, and 6.1 vs. 38%, p<0.001).

**Table 1.**
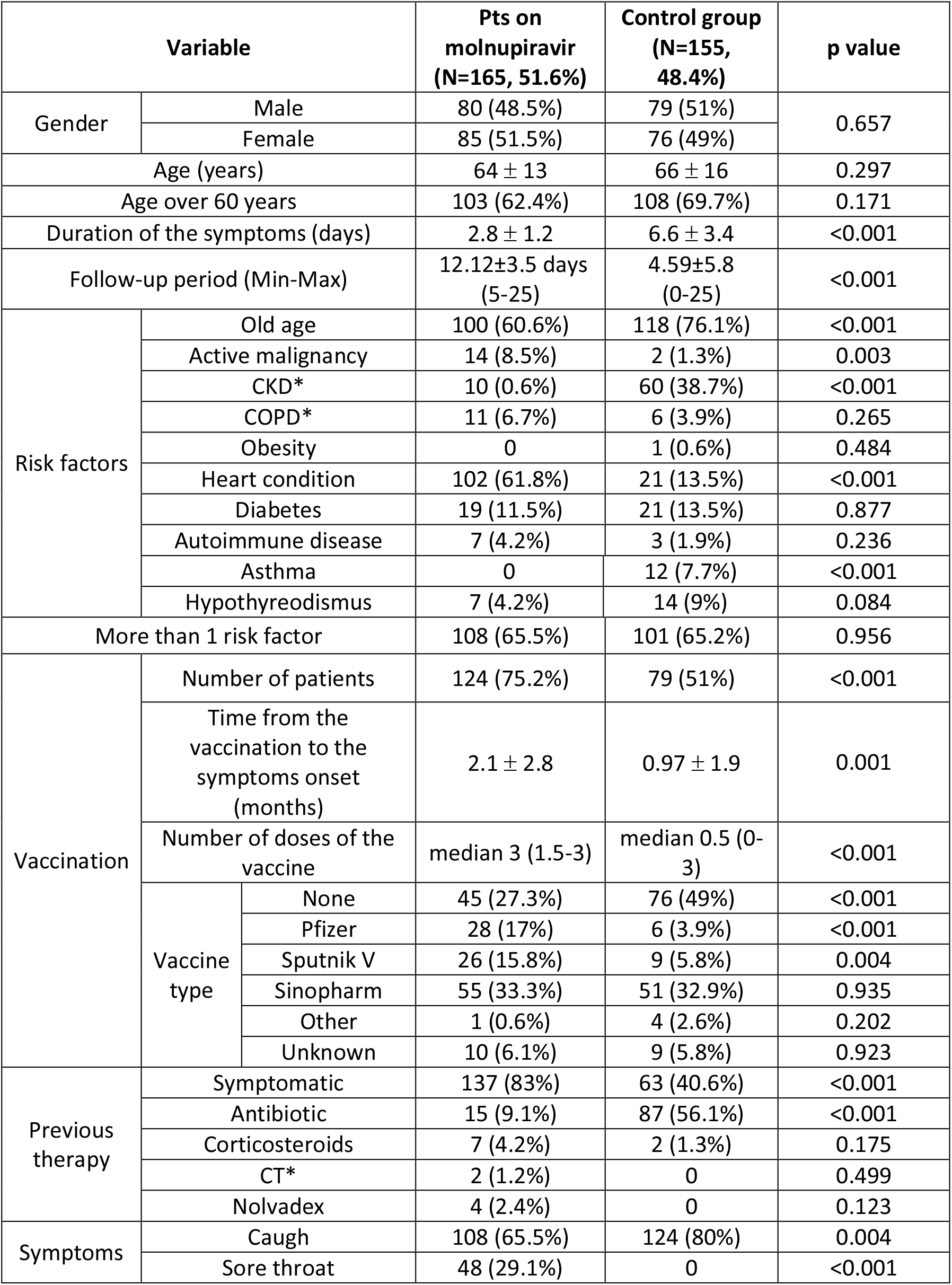
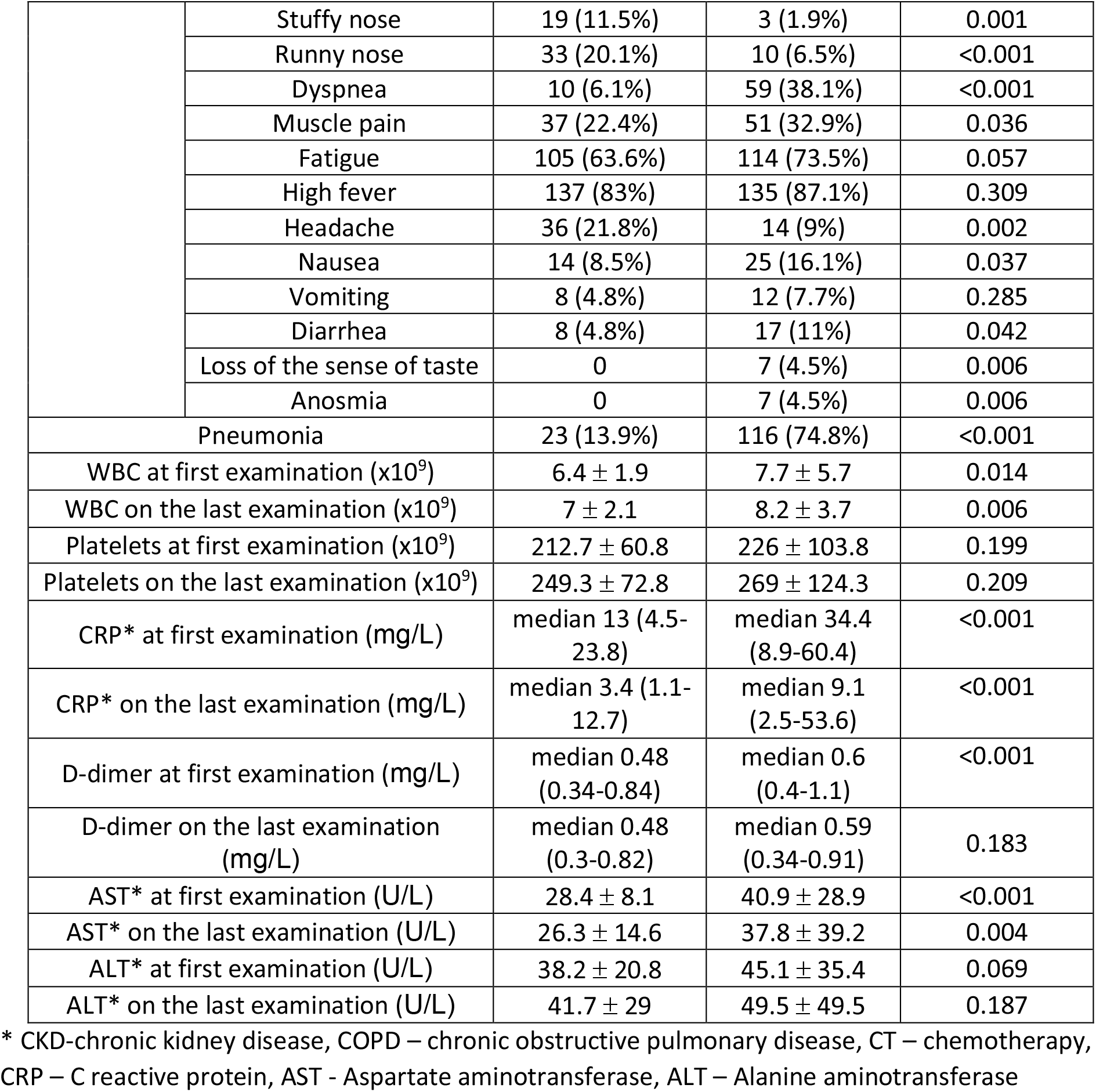
Demografic and clinical characteristics of all adult non-hospitalized patients between 15-12-2021 and 01-02-2022 receiving molnupiravir (MOL) or no antiviral therapy.

Radiographically confirmed pneumonia was predominantly experienced by patients who did not receive molnupiravir (78.4% vs 13.9%), but their symptoms also lasted longer (6.6 ± 3.4 vs 2.8 ±1.2). Regarding laboratory analysis, there was a significant difference in WBC, CRP, and AST values among groups, including measurements at the first examination compared to the last ones. Pneumonia was diagnosed in 74.8% of patients in the control group vs. 13.9%, p<0.001. (Table 1).

The majority of patients who received MOL didn’t required hospitalization (92.7 vs. 24.5%, <0.001) and the need for oxygen supplementation less frequently than those among controls (0.6% vs. 31%, <0.001). During the follow-up period of 12.12±3.5 days, none of patients on MOL were admitted to the Intensive Care Unit-ICU (vs 10.3%, <0.001). (Table 2, Figure 1). Treated patients were admitted to hospital after 8.92±3.55 days, compared to 2.83±4.95 (p<0.001) in Control group. In multivariate analysis molnupiravir was one of the independent predictors of hospitalization and reduced the possibility of hospitalization by 97.9%, while age and CRP were also independent predictors of hospitalization for an increase of one year in age, the possibility of hospitalization increased by 3.2% and for CRP by one unit 0.4%. (Table 3)

**Table 2.**
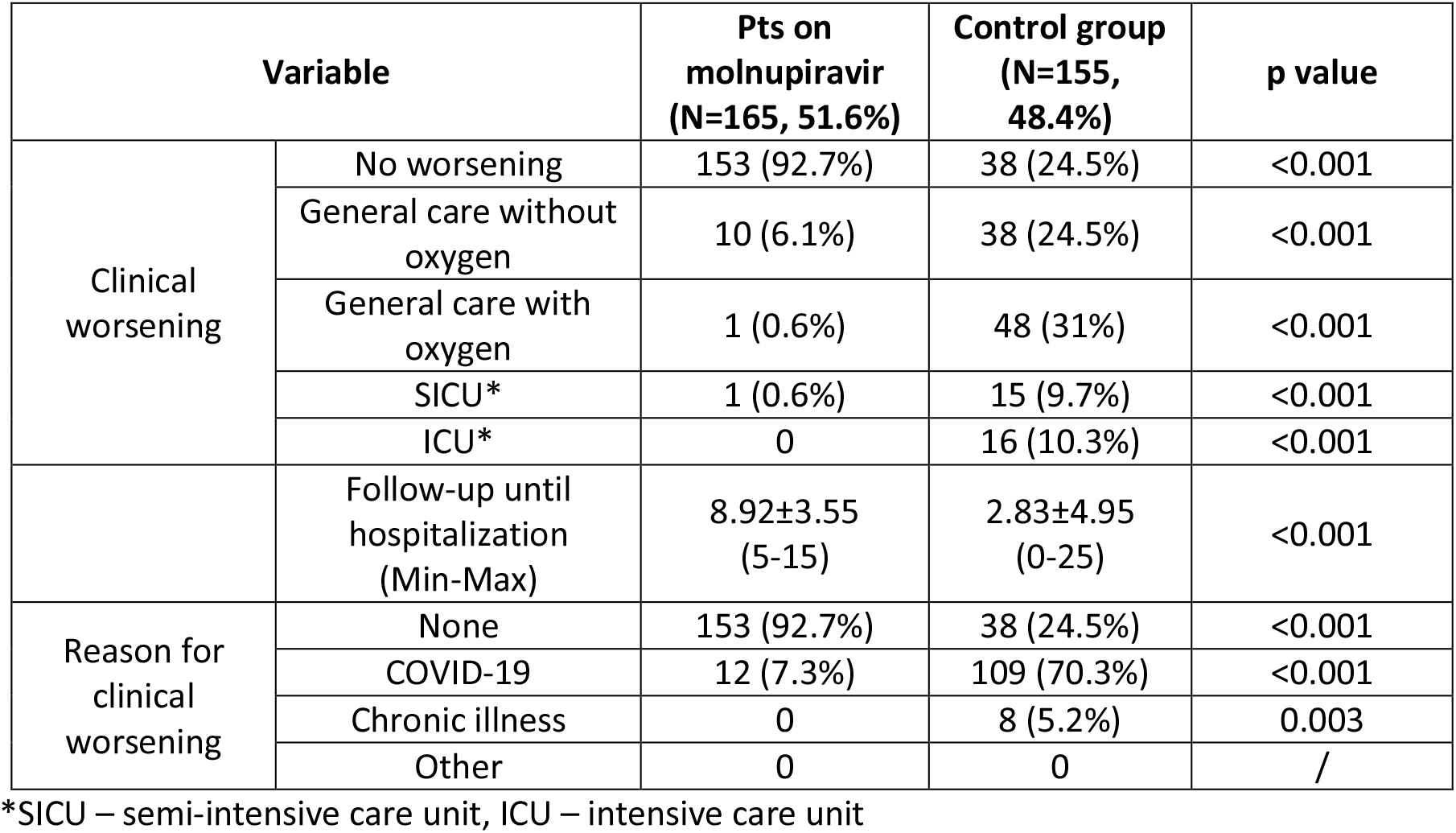
Clinical worsening and the reason for the clinical worsening between the patients with and without molnuiravir.

| Variable |  | Pts on molnupiravir (N=165, 51.6%) | Control group (N=155, 48.4%) | p value |
| --- | --- | --- | --- | --- |
| Clinical worsening | No worsening | 153 (92.7%) | 38 (24.5%) | <0.001 |
|  | General care without oxygen | 10 (6.1%) | 38 (24.5%) | <0.001 |
|  | General care with oxygen | 1 (0.6%) | 48 (31%) | <0.001 |
|  | SICU* | 1 (0.6%) | 15 (9.7%) | <0.001 |
|  | ICU* | 0 | 16 (10.3%) | <0.001 |
|  | Follow-up until hospitalization (Min-Max) | 8.92±3.55 (5-15) | 2.83±4.95 (0-25) | <0.001 |
| Reason for clinical worsening | None | 153 (92.7%) | 38 (24.5%) | <0.001 |
|  | COVID-19 | 12 (7.3%) | 109 (70.3%) | <0.001 |
|  | Chronic illness | 0 | 8 (5.2%) | 0.003 |
|  | Other | 0 | 0 | / |

\*SICU – semi-intensive care unit, ICU – intensive care unit

**Table 3.**
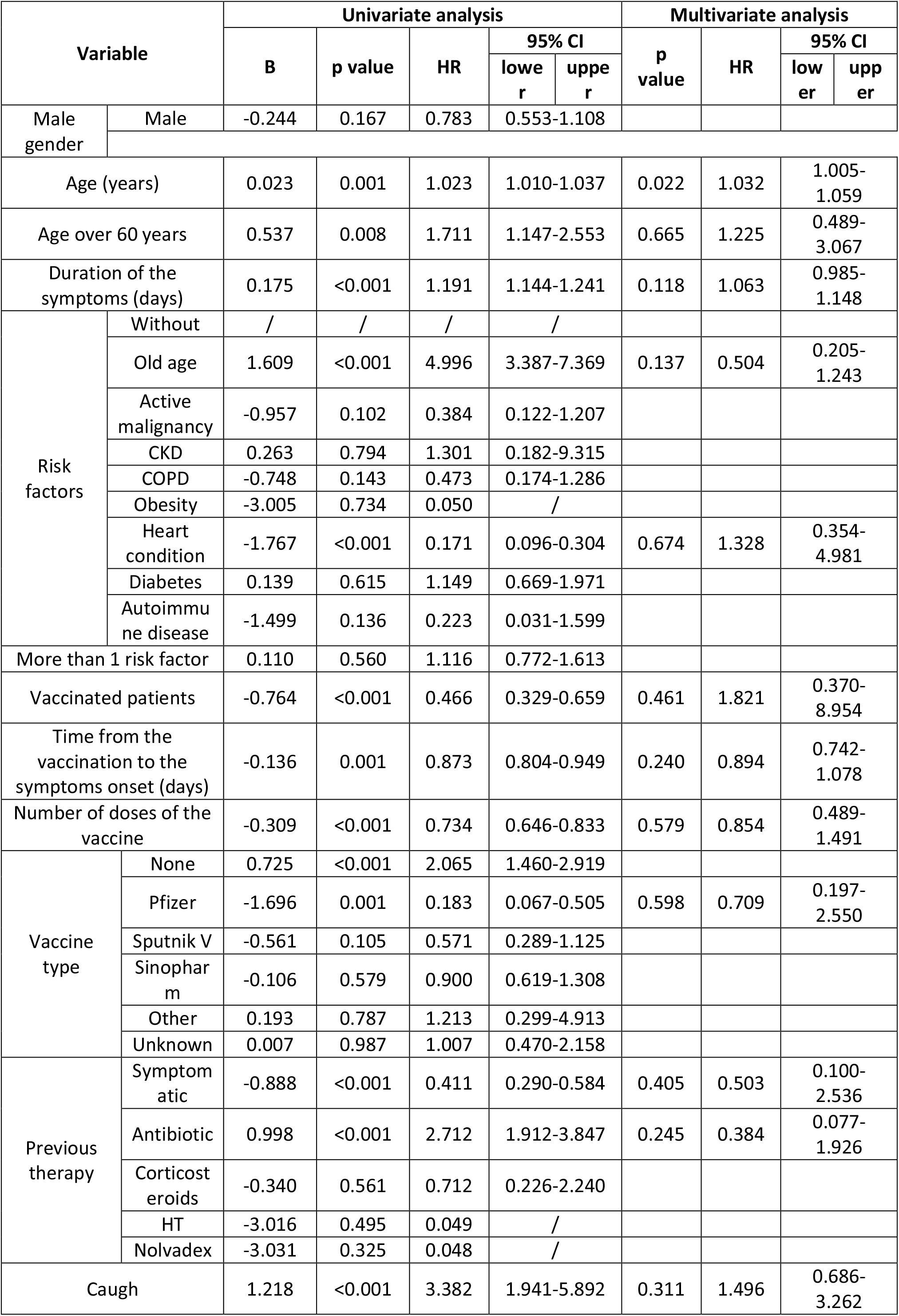
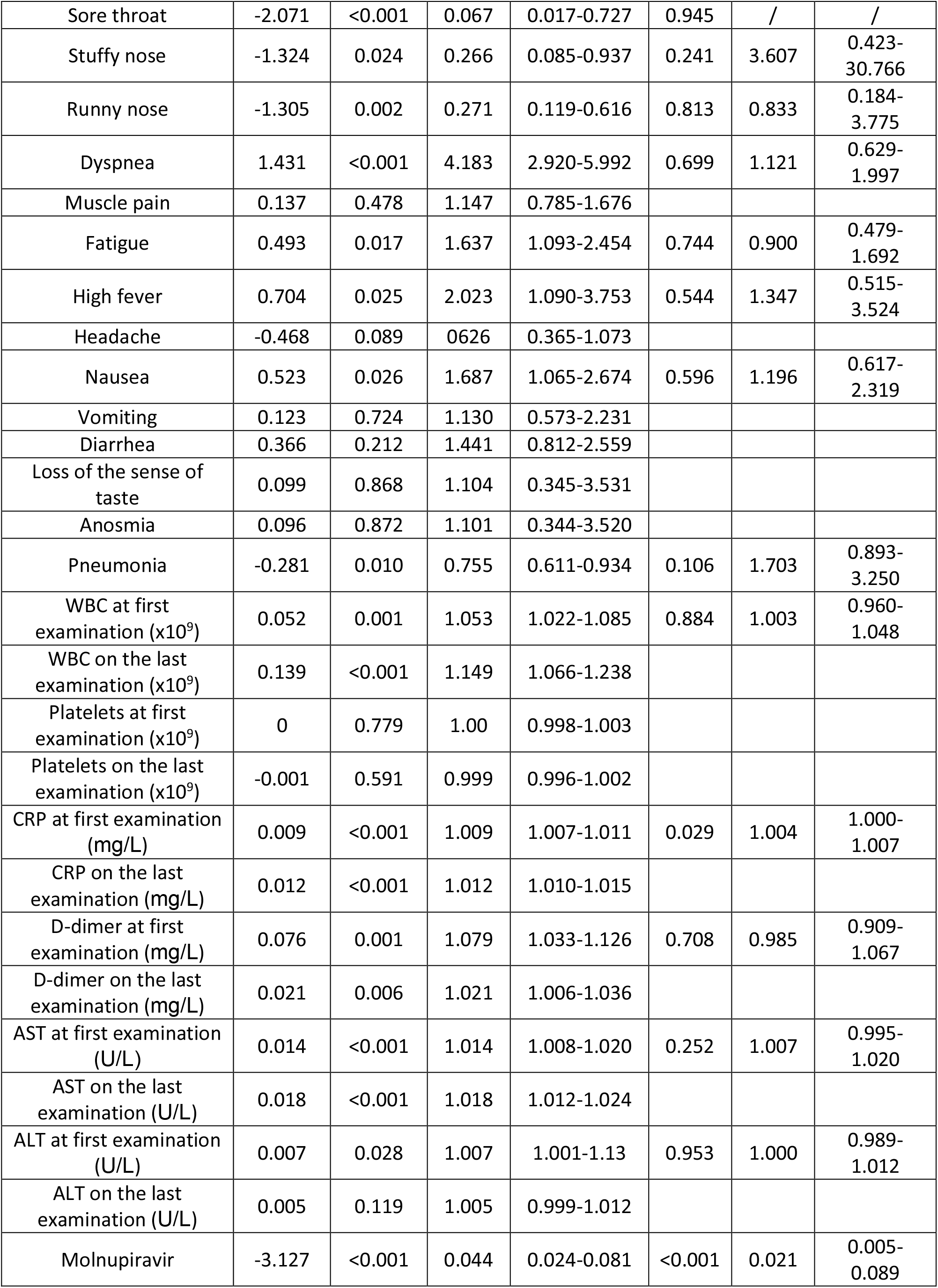
Univariate and multivariate Cox regression analyses for the hospitalization due to COVID-19

**Figure 1.**
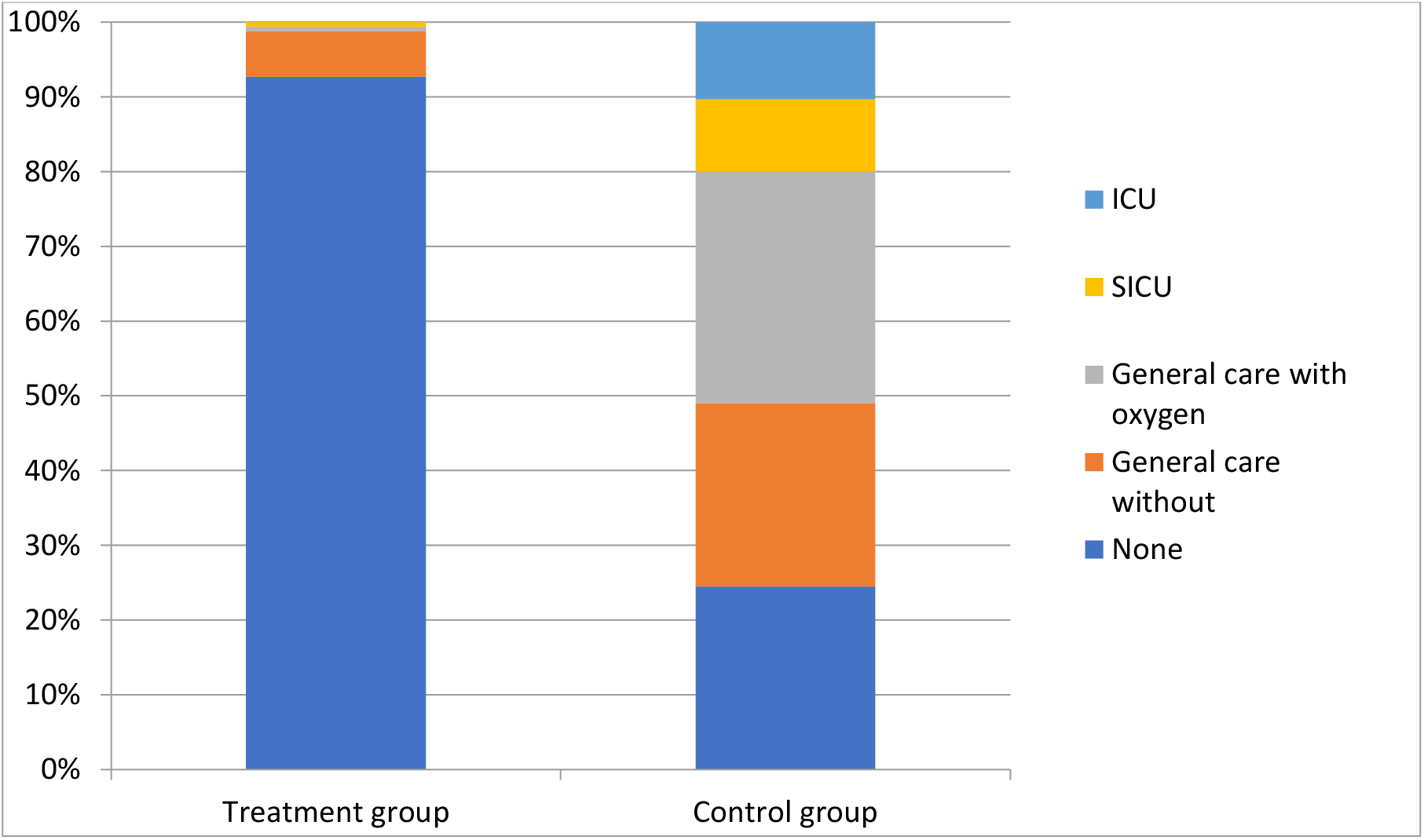

## DISCUSSION

Our study showed real-world data on the effectiveness of MOL against SARS-CoV-2 in outpatients who are at high risk of disease progression, as a single-center study. The analysis included 320 adult non-hospitalized patients treated between 12/15/2021 and 02/15/2022 in the Clinic for Infectious and Tropical Diseases, Clinical Center of Serbia, Belgrade, Serbia. Our results showed that molnupiravir reduced the risk of hospitalization (HR 0.021, p 0.001). To date, numerous studies on the effectiveness of molnupiravir showed differences from our research results; while some favored its use, others did not show greater significance.

Previous real-world effectiveness studies showed that molnupiravir use was associated with a lower risk of hospitalization in treated patients compared to non-recipients, including studies by Evans et al. (HR 0.49), Paraskevis et al. (OR 0.40), and Wai et al. (OR 0.72) [13, 14, 15]. In a study conducted among U.S. veterans aged 65 years and older, receipt of molnupiravir was associated with a lower 30-day risk of hospitalization or death (RR 0.67, 95% CI 0.46 to 0.99), but the overall result between treated and untreated patients was similar [16]. The meta-analysis by Huang et al., which included six studies involving 89,480 patients (22,355 of whom received molnupiravir therapy and 67,125 of whom received placebo therapy), indicated that the risk of mortality was reduced by 34% and the risk of the composite outcome of disease progression was reduced by 37% among patients who received molnupiravir [17].

On the other hand, Wong et al. found that molnupiravir recipients in Hong Kong had a significantly lower risk of all-cause mortality than non-recipients (HR 076 [95% CI 061-095]; p=0013), but the risk of hospitalization was comparable to the control group (098 [089-106]; p=058) [18, 19]. In another study by Yip et al., molnupiravir usage showed no statistically significant difference in reducing the risk for hospitalization versus the non-users [20]. Also, recently published 25,000 participant, prospective, open-label study, PANORAMIC trial, found only a 1% risk of all-cause hospitalization, and that molnupiravir did not reduce the frequency of COVID-19-associated hospitalizations or death among adults over 50 years of age or over 18 with other risk factors, in the community [19].

In our study, apropos that the groups were very similar in terms of demographic data, and the sole notable difference was in the number of vaccinated, there was a reason to suspect that vaccination had an impact on the more favorable course of the disease of those who were treated. That doubt has been dispelled by the fact that the Omicron variant diverged immunologically from the earlier variants on the basis of which the vaccine was evolved and with which patients may have previously come into contact. As a matter of fact, we believe that prior immune status could not have had a significant impact on the efficacy of an antiviral drug devoid of immunomodulatory activity. Our patients taking molnupiravir had a milder clinical presentation and a lower frequency of pneumonia, but they were examined earlier and started therapy on time. Based on data on follow-up time until hospitalization, regardless of the favorable course of most treated patients, they should be followed for at least 15 days after starting treatment, and those who are not on therapy should be followed for up to 25 days.

Despite the fact that it brings to our understanding of MOL use in COVID-19 patients, our study has some inherent limitations to its real-world and single-center observational nature, which might lead to bias. The main limitation of our study was the short duration of treatment as well as the small sample size. This limitation was overcome by the fact that the groups of patients proved to be similar in terms of gender and age, and this result came about by chance, without an a priori desire for matching. Because the obtained data provide significant support for expanding the base, a multicenter, national study is being planned.

Another potential limitation was the lack of data on viral subvariants.

## Data Availability

All data produced in the present study are available upon reasonable request to the authors

## CONCLUSION

We have shown that in a real-world study, the use of molnupiravir further reduced the risk for hospitalization, with the effect being more pronounced in highly vulnerable populations, suggesting that its use in high-risk populations is strongly indicated to reduce the burden of disease and unfavorable outcomes.

## References

1. WHO coronavirus (COVID-19) dashboard. Geneva: World Health Organization, 2021 (https://covid19.who.int).

2. Ko JY, Danielson ML, Town M, et al. Risk factors for coronavirus disease 2019 (COVID-19)–associated hospitalization: COVID-19–associated hospitalization surveillance network and behavioral risk factor surveillance system. Clin Infect Dis 2021;72(11):e695–e703.

3. Stokes EK, Zambrano LD, Anderson KN, et al. Coronavirus disease 2019 case surveillance — United States, January 22– May 30, 2020. MMWR Morb Mortal Wkly Rep 2020;69:759–65.

4. Kompaniyets L, Goodman AB, Belay B, et al. Body mass index and risk for COVID-19-related hospitalization, intensive care unit admission, invasive mechanical ventilation, and death — United States, March–December 2020. MMWR Morb Mortal Wkly Rep 2021;70:355–61

5. Arribas JR, Bhagani S, Lobo S, et al. Randomized trial of molnupiravir or placebo in patients hospitalized with Covid-19. NEJM Evid. DOI:.1056/EVIDoa2100044

6. Cohen MS, Wohl DA, Fischer WA, Smith DM, Eron JJ. Outpatient treatment of SARS-CoV-2 infection to prevent COVID-19 progression. Clin Infect Dis 2021;73:1717–21

7. Painter GR, Natchus MG, Cohen O, Holman W, Painter WP. Developing a direct acting, orally available antiviral agent in a pandemic: the evolution of molnupiravir as a potential treatment for COVID-19. Curr Opin Virol 2021;50:17–22.

8. Kumarasamy N, Jindal, A, Saha, B, Singh VB, Rodduturi NCR, Sinha S, Sriramadasu SC. Phase IIII trial of molnupiravir in adults with mild SARS-cov2 infection in India, 2022

9. Abdelnabi R, Foo CS, De Jonghe S, Maes P, Weynand B, Neyts J. Molnupiravir inhibits the replication of the emerging SARS-CoV-2 variants of concern (VoCs) in a hamster infection model. J Infect Dis 2021;224:749–53.

10. Gordon CJ, Tchesnokov EP, Schinazi RF, Götte M. Molnupiravir promotes SARS-CoV2 mutagenesis via the RNA template. J Biol Chem 2021;297:100770.

11. Chawla A, Cao Y, Stone J, et al. Modelbased dose selection for the phase 3 evaluation of molnupiravir (MOV) in the treatment of COVID-19 in adults. In: Proceedings and abstracts of the 31st Annual Meeting of the European Congress of Clinical Microbiology and Infectious Diseases, July 9–12, 2021. Basel, Switzerland: European Society of Clinical Microbiology and Infectious Diseases, 2021.

12. Jayk Bernal A, Gomes da Silva MM, Musungaie DB, Kovalchuk E, Gonzalez A, Delos Reyes V, et al. Molnupiravir for oral treatment of Covid-19 in nonhospitalized patients. N Engl J Med 2022;386:509–20.

13. Evans, A., Qi, C., Adebayo, J. O., Underwood, J., Coulson, J., Bailey, R., Lyons, R., Edwards, A., Cooper, A., John, G., & Akbari, A. (2023). Real-world effectiveness of molnupiravir, nirmatrelvir-ritonavir, and sotrovimab on preventing hospital admission among higher-risk patients with COVID-19 in Wales: a retrospective cohort study. The Journal of Infection. https://doi.org/10.1016/j.jinf.2023.02.012

14. Paraskevis, D., Gkova, M., Mellou, K., Gerolymatos, G., Psalida, P., Gkolfinopoulou, K., Kostaki, E. G., Loukides, S., Kotanidou, A., Skoutelis, A., Thiraios, E., Saroglou, G., Zografopoulos, D., Mossialos, E., Zaoutis, T., Gaga, M., Tsiodras, S., & Antoniadou, A. (2023). Real-world effectiveness of molnupiravir and nirmatrelvir/ritonavir among COVID-19 community, highly vaccinated patients with high risk for severe disease: Evidence that both antivirals reduce the risk for disease progression and death. https://doi.org/10.1101/2023.02.09.23285737

15. Wai, A. K.-C., Chan, C. Y., Cheung, A. W.-L., Wang, K., Chan, S. C.-L., Lee, T. T.-L., Luk, L. Y.-F., Yip, E. T.-F., Ho, J. W.-K., Tsui, O. W.-K., Cheung, K. W.-Y., Lee, S., Tong, C.-K., Yamamoto, T., Rainer, T. H., & Wong, E. L.-Y. (2023). Association of Molnupiravir and Nirmatrelvir-Ritonavir with preventable mortality, hospital admissions and related avoidable healthcare system cost among high-risk patients with mild to moderate COVID-19. The Lancet Regional Health. Western Pacific, 30(100602), 100602. https://doi.org/10.1016/j.lanwpc.2022.100602

16. Bajema, K. L., Berry, K., Streja, E., Rajeevan, N., Li, Y., Yan, L., Cunningham, F., Hynes, D. M., Rowneki, M., Bohnert, A., Boyko, E. J., Iwashyna, T. J., Maciejewski, M. L., Osborne, T. F., Viglianti, E. M., Aslan, M., Huang, G. D., & Ioannou, G. N. (2022). Effectiveness of COVID-19 treatment with nirmatrelvir-ritonavir or molnupiravir among U.S. Veterans: target trial emulation studies with one-month and six-month outcomes. MedRxiv: The Preprint Server for Health Sciences. https://doi.org/10.1101/2022.12.05.22283134

17. Huang, C., Lu, T.-L., & Lin, L. (2023). Real-world clinical outcomes of molnupiravir for the treatment of mild to moderate COVID-19 in adult patients during the dominance of the Omicron variant: A meta-analysis. Antibiotics (Basel, Switzerland), 12(2), 393. https://doi.org/10.3390/antibiotics12020393

18. Wong, C. K. H., Au, I. C. H., Lau, K. T. K., Lau, E. H. Y., Cowling, B. J., & Leung, G. M. (2022). Real-world effectiveness of molnupiravir and nirmatrelvir plus ritonavir against mortality, hospitalisation, and in-hospital outcomes among community-dwelling, ambulatory patients with confirmed SARS-CoV-2 infection during the omicron wave in Hong Kong: an observational study. Lancet, 400(10359), 1213–1222. https://doi.org/10.1016/S0140-6736(22)01586-0

19. Butler, C. C., Hobbs, F. D. R., Gbinigie, O. A., Rahman, N. M., Hayward, G., Richards, D. B., Dorward, J., Lowe, D. M., Standing, J. F., Breuer, J., Khoo, S., Petrou, S., Hood, K., Nguyen-Van-Tam, J. S., Patel, M. G., Saville, B. R., Marion, J., Ogburn, E., Allen, J., … PANORAMIC Trial Collaborative Group. (2023). Molnupiravir plus usual care versus usual care alone as early treatment for adults with COVID-19 at increased risk of adverse outcomes (PANORAMIC): an open-label, platform-adaptive randomised controlled trial. Lancet, 401(10373), 281–293. https://doi.org/10.1016/S0140-6736(22)02597-1

20. Yip, T. C.-F., Lui, G. C.-Y., Lai, M. S.-M., Wong, V. W.-S., Tse, Y.-K., Ma, B. H.-M., Hui, E., Leung, M. K. W., Chan, H. L.-Y., Hui, D. S.-C., & Wong, G. L.-H. (2023). Impact of the use of oral antiviral agents on the risk of hospitalization in community Coronavirus disease 2019 patients (COVID-19). Clinical Infectious Diseases: An Official Publication of the Infectious Diseases Society of America, 76(3), e26–e33. https://doi.org/10.1093/cid/ciac687

